# Continuous Positive Airway Pressure (CPAP) in Covid 19 Acute Respiratory Distress Syndrome (ARDS): A Systematic Review

**DOI:** 10.1101/2022.11.29.22282819

**Authors:** Anbesan Hoole

**Affiliations:** Respiratory Physician, Bach Christian Hospital, Qalandarabad, Abbottabad District, KPK, Pakistan

## Abstract

**Introduction:** Acute Respiratory Distress Syndrome (ARDS) is a feared consequence of Covid 19 Pneumonia. Traditional guidance was for ARDS to be treated with Intubation and Mechanical Ventilation (IMV), when failing simple oxygen. However globally numbers of patients with Covid 19 ARDS (CARDS) quickly overwhelmed IMV capacity, with Continuous Positive Airway Pressure (CPAP) has been used as a bridge or alternative to IMV. However, the evidence base remains limited in quality despite widespread adoption in guidelines.

**Methods:** Pubmed (15.6.2022), Embase (30.7.2022) and Google Scholar (4.8.2022) were searched to identify studies with the primary outcome of IMV free survival in patients with CARDS receiving CPAP, ideally with simple oxygen as a comparator. Secondary outcomes were overall survival with CPAP, length of stay and adverse events. All studies were assessed by the relevant Critical Appraisal Skills Programme Tool (CASP).

**Results:** 13 studies were identified, out of which only 1 was a Randomised Control Trial (RCT) with simple oxygen as a comparator. There were 11 Cohort studies and one Systematic review.

**Discussion:** There is much heterogeneity in CPAP success rates (50 – 70%), which may be linked to variation in candidate selection, resource setting, application protocols and combined use with other respiratory support modalities (Non Invasive Ventilation – NIV, and High Flow Nasal Oxygen – HFNO). Adverse events and economic data such as length of stay are under reported.

**Conclusion:** CPAP is an effective respiratory support in CARDS particularly in resource poor settings. However further research is needed to refine optimum candidate selection, application protocols and any added benefit from combination with NIV or HFNO.

No funding was received for this study. This review was not registered.

## 1. Introduction

The Severe Acute Respiratory Syndrome Coronavirus 2 (SARS-CoV-2) or Covid 19 pandemic has been one of the most important public health concerns of the past century, with significant challenges regarding infection prevention and control as well as treatment of individuals with Covid 19 disease. Around 15% of unvaccinated individuals develop a severe pneumonia requiring hospitalisation for hypoxia. Out of these patients with severe pneumonia roughly one third (5% total) develop critical disease or Covid 19 Acute Respiratory Distress Syndrome (Covid 19 ARDS or CARDS) with a mortality rate of 30 – 60%(1,2).

ARDS is a feared consequence of many infective or inflammatory processes and is thought to involve leakage of fluid from blood vessels throughout the lung resulting air sacs (alveoli) filling up with fluid and severely impairing oxygen exchange. ARDS is defined by Berlin criteria as (i) acute onset (within 7 days) of (ii) hypoxia with (iii) bilateral infiltrates on chest radiograph or CT, (iv) in the presence of preserved cardiac function. ARDS is further graded as mild (P_a_O_2_/FiO_2_ 200 – 300mmHg), moderate (P_a_O_2_/FiO_2_ 100 – 200mmHg) or severe (P_a_O_2_/FiO_2_ < 100mmHg) according to the degree of hypoxia despite a Positive End Expiratory Pressure (PEEP) of 5cm H_2_O(3). However, ARDS according to this definition is difficult to demonstrate in many resource poor settings due to the requirement for PEEP usually delivered by Intubation and Mechanical Ventilation (IMV) which is in short supply. Therefore, the Berlin definition has been adapted for resource limited settings in the form of the Kigali Criteria which removes the requirement for PEEP as part of the definition(4). Historically treatment for ARDS has been supportive with IMV for patients failing simple oxygen support(5). However, IMV requires general anaesthesia and is expensive, resource intensive and impractical outside the setting of intensive care units (ICUs). The advent of Continuous positive airway pressure (CPAP) and Non-Invasive Ventilation (NIV, also known as Bilevel Positive Airway Pressure – BiPAP) devices which are less invasive, less expensive, less resource intensive and practicable outside ICUs has created scope for an alternative approach to respiratory support in ARDS.

However, application of CPAP or NIV as respiratory support either as a bridge or alternative to IMV in ARDS had limited evidence in the pre Covid 19 era, and the use of CPAP for ARDS was generally discouraged in line with a previous landmark Randomised Control Trial (RCT)(6). As such initial guidance for management of Covid 19 ARDS or CARDS focussed purely on IMV in ICU for patients failing simple oxygen(7). However, the sheer number of patients with CARDS created a pandemic of ARDS unprecedented in the modern era, overwhelming the IMV and ICU capacity of even resource rich settings(8). Resource poor settings struggled even more, with many patients struggling to receive even simple oxygen yet alone any form of respiratory support(9). Within this context, CPAP and to a lesser extent NIV were trialled as alternative respiratory support for CARDS out of necessity in Italy and other European countries early in the pandemic with promising results(10,11). This challenged the previous dogma around CPAP in ARDS(12). Even so, towards the end of the pandemic major guidelines began advocating the use of CPAP for CARDS(13,14). This systematic review seeks to explore the evidence base for CPAP as a bridge or alternative to IMV in CARDS. The primary outcome under investigation is IMV free survival in patients with CARDS treated with CPAP, ideally with simple oxygen as a comparator. Secondary outcomes include overall survival with CPAP, length of stay as a patient outcome and economic indicator, as well as any adverse events associated with CPAP therapy.

Although CARDS is decreasing globally, population growth and increasing globalisation will continue to increase the likelihood similar viral pandemic ARDS in the future(15). The question is not whether this may occur again but when. Improving our understanding regarding the utility of CPAP in CARDS will help health systems be better prepared to face future viral pandemics with ARDS, particularly those in resource poor settings which lack ICU and IMV capacity(16). Further, it may also help improve management of ARDS related to non-Covid 19 conditions.

## 2. Methods

The research question was formulated into a PICO format and used to search available medical literature on the Pubmed (Medline) and Embase databases and a Google Scholar search was conducted.

The PICO question was as follows:

- Population (P): CARDS
- Intervention (I): CPAP
- (Comparison (C): Simple oxygen)
- Outcome (O): CPAP success rate (IMV free survival)

The comparison of simple oxygen was desired for studies but not required due to limited evidence base.

Other secondary outcomes of interest were overall survival rate, length of stay (LOS) in hospital, and any adverse effects of CPAP.

The inclusion criterion was the reporting of CPAP specific intubation free survival rates in Covid 19 ARDS.

Exclusion criteria were as follows

1. Articles where CPAP specific intubation free survival rates were not reported as separate from other forms of Non-invasive Respiratory Support (NRS, i.e., NIV and High Flow Nasal Oxygen – HFNO)
2. Articles which did not report survival data until at least 28 days
3. Articles without full length English Manuscripts
4. Articles focussing exclusively on special patient groups (e.g., pregnancy, renal failure).

Pubmed, Embase and Google scholar were selected for literature search, and article selection was completed by the author without any automation of the process. All selected articles were analysed using the relevant Critical Appraisal Skills Programme (CASP) checklist. No meta-analysis was conducted due to heterogeneity of studies.

Patients and public were not involved in the design, conduct, reporting or dissemination of research plans.

## 3. Results

### Study selection

Details of the Pubmed, Embase and Google Scholar searches are given below (Table 1):

**Table 1:** Details of searches of Pubmed, Google Scholar and Embase data bases

| Data base | Date | Search terms | Results |
| --- | --- | --- | --- |
| Pubmed | 15.6.2022 | *(Covid OR sars coronavirus OR sars-cov-2) AND (acute respiratory distress syndrome OR ards OR severe pneumonia) AND (continuous positive airway pressure OR cpap)* | • 198 results for screening<br>• 72 records sought for retrieval<br>• 10 after exclusion criteria applied |
| Google scholar | 30.7.2022 | *Covid OR sars coronavirus OR sars-cov-2) AND (acute respiratory distress syndrome OR ards OR severe pneumonia) AND (continuous positive airway pressure OR cpap)* | • Yielded 15 400 results in total<br>• First 100 selected by relevance for screening<br>• 22 sought for retrieval including after removal of duplicates from prior searches<br>• 2 additional full length articles included after exclusion criteria applied |
| Embase | 4.8.2022 | *(Covid OR sars coronavirus OR sars-cov-2) AND (acute respiratory distress syndrome OR ards OR severe pneumonia) AND (continuous positive airway pressure OR cpap)* | • Yielded 171 results in total<br>• 8 sought for retrieval including after removal of duplicates.<br>• 1 additional full-length article included after exclusion criteria applied. |

A total of 13 articles (Cohort study (n = 11), Randomised Control Study (n = 1) or Systematic Review (n = 1)) were selected for Systematic Review after the above inclusion and exclusion criteria were applied, as seen in the PRISMA flow diagram(17) below (Figure 1).

**Figure 1:**
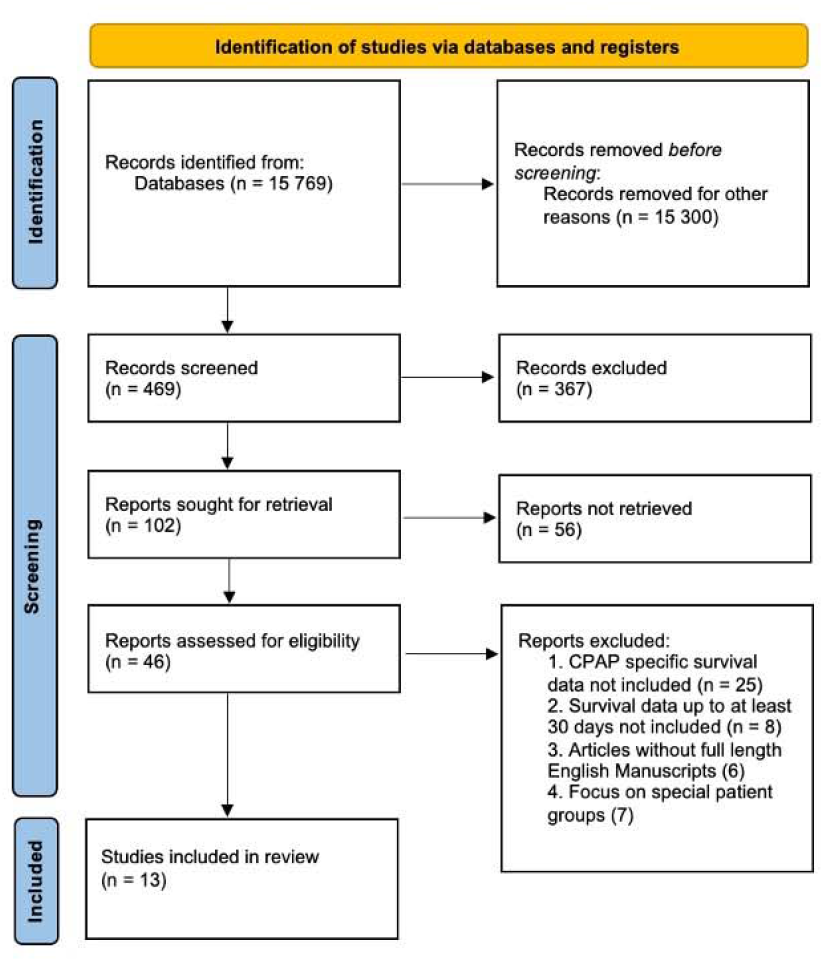
PRISMA flow diagram for article selection for critical review

### Studies for review

Individual CASP analyses for the 13 articles selected can be found in supplement 1. A summary of the 13 studies for CASP analysis is presented below (Table 2):

**Table 2:** Summary of the studies included for Critical Appraisal on CPAP in CARDS

| Study | Population | Intervention | Comparison | Outcome |
| --- | --- | --- | --- | --- |
| 1. Brusasco et al (2021)(19)<br>Observational Study:<br>Prospective Cohort<br>Single Centre (Genoa, Italy).<br>Sample size N=64 | Covid 19 ARDS (Moderate to Severe by Berlin Criteria) | CPAP<br>(n = 64) | None | 83% success rate with CPAP<br>53/64 (Survive with CPAP)<br>4/64 (Died under CPAP)<br>7/64 received IMV (2 survived, 5 died) |
| 2. Aliberti et al (2020)(20)<br>Multicentre prospective<br>cohort study (Milan, Italy)<br>N = 157 | Covid 19 ARDS (Mild to Severe) | Helmet CPAP (n = 157, 2 centres) | None | 55% success rate with CPAP<br>87/157 (Survive with CPAP)<br>36/157 (Died under CPAP)<br>34/157 received IMV (25 survived, 9 died) |
| 3. Franco et al (2020)(21)<br>Multicentre prospective<br>cohort 4 hospitals (Italy)<br>N = 670 | Covid 19 ARDS (Mild to Severe) | CPAP (330) | HFNC (163)<br>and NIV (177) | 70% success rate with CPAP<br>230/330 (survive on CPAP)<br>74/330 (Died under CPAP)<br>82 received IMV (58 survive, 24 died) |
| 4. Radovanovic et al (2021)<br>(22) Systematic Review: 23<br>studies published between<br>Dec 2019 and Nov 2021 | Covid 19 ARDS (Mild to Severe) | CPAP or NIV | None | Success 51.3% in CPAP<br>Success 41.2% in NIV |
| 5. Hoole et al (2022)(23)<br>Single centre retrospective<br>cohort study. N = 99 | Covid 19 ARDS (Moderate to Severe) | Facemask CPAP | None | 76.8% success and survival rate on CPAP for Moderate to Severe Covid 19 ARDS (76/99). 5% Barotrauma rate. No escalation to IMV due to resource scarcity. |
| 6. Coppadoro et al (2021)(24)<br>Single centre retrospective<br>cohort study. N = 306 | Covid 19 ARDS (Mild to Severe) | Helmet CPAP | None | 52% CPAP success rate (28% in frail patients with do not intubate (DNI) orders, 69% in full treatment group). Patients with CPAP success significantly more likely to show a doubling in PaO<sub>2</sub>/FiO<sub>2</sub> with CPAP, compared with CPAP failures. |
| 7. Isaac et al. (2022)(25)<br>Single centre retrospective<br>cohort study. N = 168 | Covid 19 ARDS (Moderate) | Facemask CPAP and NIV in case of CPAP failure | None | 41% (69/168) success rate with CPAP alone, 36% (60/168) additional success rate with CPAP followed by NIV. Overall CPAP + NIV success rate of 77% (129/168). Of those failing CPAP + NIV and for full escalation, 3/9 receiving IMV survived. |
| 8. Coppola et al (2021)(26)<br>Retrospective multi centres<br>(2 hospitals) observational<br>cohort study. N = 156 | Covid 19 ARDS (Moderate to Severe) | Helmet CPAP + NIV trial in case of CPAP failure if not immediately needing IMV | None | CPAP successful in 93/156 (60%), CPAP + CPAP/NIV successful in 109/156 (70%). 20/47 receiving IMV survived (43%). Overall survival 129/156 (83%) |
| 9. Ashish et al (2020)(27).<br>Single centre two period<br>retrospect comparison<br>cohort study. N = 206 | Covid 19 ARDS | Early CPAP (< 7 days<br>from admission) | Late CPAP (>7<br>days from<br>admission) | Early CPAP protocol associated with an increased survival<br>(8/15 in late group, 1/3 in early group) |
| 10. Wozniak et al (2022)(28).<br>Single centre retrospective<br>cohort study. N = 23 | Covid 19 ARDS | CPAP | None | 14/23 (61%) responded to CPAP. 9/9 Non responders<br>survived on IMV after failed CPAP trial. |
| 11. Perkins et al (2022)(29).<br>Multicentre Adaptive RCT (N<br>= 1273) | Covid 19 ARDS (Moderate to<br>Severe) | CPAP or HFNO | Simple<br>oxygen | 240/377 (64%) intubation free survival at 30 days in CPAP<br>group vs 198/356 (56%) in simple oxygen (p = 0.03);<br>231/415 (56%) in HFNO vs 202/368 (55%) in simple<br>oxygen |
| 12. Vaschetto et al<br>(2020)(30). Single centre<br>retrospective cohort study (N<br>= 537) | Covid 19 ARDS (Moderate to<br>Severe) | CPAP | None | 55% intubation free survival with CPAP and 66% survival<br>overall (79% in full escalation group and 27% in DNI<br>group). Delayed intubation associated with increased<br>mortality. |
| 13. Lawton et al (2021)(31).<br>Single centre retrospective<br>cohort study (N = 165) | Covid 19 ARDS (Moderate to<br>Severe) | CPAP | None | 62% intubation free survival with CPAP and early proning<br>in full escalation group. |

## 4. Discussion

The utility of CPAP in Covid 19 ARDS (CARDS) as opposed to simple oxygen before IMV has been an area of dynamic growth in knowledge and understanding. Early observational studies utilising CPAP in CARDS arose out of a pragmatic approach to respiratory support in CARDS as availability of IMV quickly outstripped the demand in Italian hospitals. Since then, studies have explored the adoption of CPAP, NIV and HFNO before IMV in a wide variety of settings using differing protocols.

13 studies were identified in this study using Medline (Pubmed), Embase and Google Scholar. These data bases were selected for their coverage of medical literature. Limitations of this search methodology include exclusion of unpublished literature and literature in other languages. Cascading citation expansion could have been utilised as an incremental method to find additional articles but was not utilised. Further there is likely a publication bias in centres with better CPAP outcomes being more likely to publish their data than those with poorer, However, these studies still provided a broad overview of CPAP success (intubation free survival) in a wide variety of settings.

An analysis of the above studies can be explored along the following themes

### 1) Type of study: Observational vs Randomised Control Trial (RCT)

Most studies (11/13) were observational with no clear comparison to simple oxygen which is a significant limiting factor. While these observational studies can provide some supportive evidence for the use of CPAP in CARDS, they lack the strength of interventional studies in terms of the degree of evidence provided. Even so, they were significant first steps in the context of a new and deadly pandemic disease supporting the use of CPAP in CARDS.

One study (Perkins et al) was an adaptive RCT, which provided a better level of evidence for CPAP in CARDS and shaped national guidelines in the UK as mentioned before.

One study was a Systematic Review, with a CPAP success rate of 51.3%. However, since some of the data came from studies including mild ARDS, it is unlikely that CPAP avoided intubation in all cases of success. Further all the studies included were observational cohort studies, and therefore inference of treatment success to CPAP difficult as mentioned before. Of note, CPAP success rate was higher than NIV (41.2%) but it is unclear if the difference is significant due to lack of confidence intervals.

### 2) Early versus Late studies

Studies within the first few months of the Covid 19 pandemic (Early 2020) reported high overall CARDS mortality rates and high CPAP failure rates(11,33). However, studies later in the pandemic reported lower overall CARDS mortality rates and lower CPAP failure rates(10,23,29). Interestingly there is also a pre Covid 19 study from Southern China reporting good outcomes with CPAP in SARS related ARDS(34) but further exploration of this is not the topic of this research.

Reasons improving CARDS mortality and better CPAP outcomes over time are unclear but could reflect better treatment for CARDS over time as experience and evidence amounted, and possibly decreased virulence of the virus. This is a significant confounder in the two-period study by Ashish et al comparing earlier management with simple oxygen against later studies with CPAP. While an improvement in outcomes was seen with CPAP, the time difference is a significant confounder for the reasons mentioned before.

### 3) Patient cohort selection: Full escalation versus Do Not Intubate (DNI) patients

In general, elderly patients with multiple comorbidities worse outcomes with CARDS and are often subject to DNI status on hospitalisation, as opposed to patients with a better functional baseline who are for full escalation and may be considered for IMV on failure of simple oxygen or Non-Invasive Respiratory Support (NRS). Where patients were segregated into DNI and full escalation groups, DNI patients had a CPAP success rate of around 30%, whereas as full escalation patients had CPAP success rates of 70 – 80% or more. Overall CPAP success rates were generally around 50 – 70% (Range 41% - 83%). Variation in cohort age and comorbidity demographics likely contributes to a significant amount of the variation in CPAP in success rates observed.

The difference in CPAP performance between DNI and full escalation patients raises a dilemma in settings severely constrained by staff and equipment resources. On the one hand, 3/10 DNI patients with CARDS may have a survival benefit with CPAP. However, on the other hand DNI patients are much less likely to have a successful outcome with CPAP than full escalation patients. Therefore, a pragmatic approach may be to provide DNI patients with CPAP when equipment and staff are available, but to prioritise full escalation patients when resources are severely constrained.

### 4) Location of study: Higher Income Countries (HICs) versus LMICs

Most of the studies of CPAP in CARDS come from High Income Countries (Italy – 6, UK – 4). There are two from LMICs (India – 1, Pakistan - 1). Regarding resource limited settings, the latter two studies from mission hospitals in Pakistan and India suggest CPAP can be used in CARDS with good outcomes. However, a study not included in this review from Baghdad suggests caution. However, this may be due to heterogeneity in CPAP application protocols, as well as staff and patient confidence.

### 5) Heterogeneity in CPAP application protocols

While consensus criteria for some aspects of CPAP application emerged later on in the pandemic in the form of guidelines (e.g. NICE), there was much heterogeneity in the use of CPAP for most of the pandemic. This is reflected in these studies with variation of the following aspects.

#### I. Initiation of CPAP

Most studies-initiated CPAP during moderate to severe ARDS, while in some studies CPAP was initiated even in mild patients. In many studies an FiO2 requirement of 0.4 or more to maintain oxygenation (corresponding to moderate to severe ARDS by P_a_O_2_/FiO_2_ ratio) was considered an indication for CPAP and this is reflected in the NICE guidelines. Use of CPAP in mild disease in a study without non-CPAP controls may be expected to increase the success rate of CPAP measured as survival or intubation free survival. However, as these patients may have survived without intubation even without CPAP, it is unclear if this is truly representing CPAP success.

Conversely, delaying CPAP initiation until severe ARDS may decrease CPAP success rates not only because of the disease severity of selected patients, but also because the optimum time window for intervention is lost(30). In the early phase of Covid 19 ARDS the lungs appear to have a higher compliance with greater alveolar recruitability with CPAP, whereas later with progression of CARDS compliance is reduced and alveolar recruitability is lost(35,36). This is shown in studies showing greater CPAP success rate and survival with early compared with late CPAP(27). Further one of the mechanisms by which CPAP is postulated to improve outcomes in ARDS is through reduction of patient self-inflicted lung injury (P – SILI) through preventing hyperventilation(37). Again, delaying CPAP increases the amount of cumulative injury through reduction of time spent in protective ventilation.

#### II. CPAP machine and interface

Many of the early Italian trials utilised helmet CPAP due to concerns regarding aerosolization, as opposed to facemask CPAP used in other areas later in the pandemic. While this represents a difference in the CPAP set up, the impact on CPAP efficacy is unclear.

#### III. Use of prone positioning

Awake prone positioning has been shown to improve outcomes in CARDS, with a landmark RCT reporting a Hazard Ratio of 0.75 for IMV and 0.87 for 28-day mortality(38). Awake prone positioning with helmet or facemask CPAP was adopted variably through the studies. Notably some of the studies with a higher CPAP success rate adopted awake prone positioning routinely as part of their protocol, although for differing durations.

#### IV. Definition of CPAP failure

There is significant heterogeneity around the definition of CPAP failure. In general, earlier studies in resource rich settings feature early escalation to intubation and ventilation based on parameters such as raised respiratory rate whereas later studies and those in resource poor settings feature continued CPAP despite raised respiratory rate which was shown to be a poor marker of CPAP failure(25,28). This remains a significant area of need for further research. Further while greater duration on CPAP appears to be associated with poorer outcomes, it clear that some patients appear to show good tolerance to longer duration of CPAP with better survival outcomes than initially anticipated. Further research is needed to define this patient cohort. CPAP failure due to patient intolerance also needs to be considered, as well as the possible role of breaks with HFNO and judicious use of sedatives to improve compliance. Further research is needed regarding the optimal use of these adjuncts.

#### V. Weaning plan for CPAP

Again, there is much variation around the weaning protocol for CPAP, with rapid versus slow weans and pressure versus time based weans. Optimal protocols for CPAP weaning need to be investigated further.

### 6) CPAP alone versus CPAP plus other Non-Invasive Respiratory Support Modalities (NIV and HFNO)

#### I. NIV

Two studies in this review (Isaac et al and Coppola et al) advocate for the use of NIV as a step-up modality for patients failing CPAP, reporting 36% and 10% additional success rates respectively with NIV added to CPAP. However, in the case of Isaac et al patients were deemed to be CPAP failures if there was insufficient improvement within 30 min of CPAP initiation which would seem early given British Thoracic Society (BTS) standards advocating at least an hour with NRS(39), and in both cases CPAP pressures higher than 10cm H2O were not trialled as in some other studies(23,31).

Further given that Covid 19 ARDS in general presents with type I respiratory failure rather than type 2 respiratory failure representing a ventilatory defect(40), the general use of NIV for patients failing CPAP would seem questionable, particularly without the demonstration of type 2 respiratory failure by arterial blood gas (ABG).

However, NIV may certainly benefit a specific group of patients failing CPAP, particularly in the context of limited access to IMV. Additionally, a minority of patients present with type 2 respiratory failure in CARDS, often due to underlying comorbidities such as Chronic Obstructive Pulmonary Disease (COPD), and these patients are also likely to benefit from NIV(40). Further research is needed, to define these two patient cohorts who may benefit from NIV, and the characteristics of the NIV protocols to be adopted. However, routine use of NIV as first line appears to have inferior outcomes to CPAP from the study of Radanovic et al. But further exploration of the use of NIV in this context is not the subject of this paper.

#### II. HFNO

As mentioned before, HFNO has often been used to provide breaks for patients on CPAP and may improve compliance. However, no study explored HFNO as a step up for patients failing CPAP and given the PEEP requirement of these CARDS patients to maintain oxygenation, HFNO is unlikely to play an additional role. Further HFNO alone was not shown superior to simple oxygen in the Perkins et al (Recovery RS) trial, but this may be due to the trial being under powered to detect a difference (type 2 error)

### 7) Adverse Effects and Long-term effects

In general, adverse effects of CPAP were under reported but includes barotrauma(41). Even so rates of barotrauma may be less with CPAP than with IMV. Long term outcomes of CARDS treated with CPAP appear good and non-inferior to early IMV(42).

### 8) Data for Economic Analyses

Data regarding length of stay was under reported, and no study included information on costs. These are crucial for cost benefit or cost utility analyses to make future funding decisions on CPAP for CARDS or other similar pandemic viral ARDS. Economic analyses are essential alongside clinical analyses, to allow local and national authorities to make decisions on public health policy. This is an essential topic for future research.

## 5. Conclusion

### Main outcome

While the heterogeneity in the application of CPAP and target patient cohorts makes a generalised estimation of CPAP success rates difficult, most studies give success rates of 50 – 70%, meaning that in 50 – 70% of cases CPAP can avoid IMV. This makes CPAP an effective management strategy in CARDS, particularly within a context of limited resources. While some studies advised avoidance of CPAP in severe ARDS, a few studies suggests that in even in severe ARDS a trial of CPAP can be attempted with good outcomes. The largest RCT to date (Recovery RS) shows intubation free survival benefit of CPAP over simple oxygen which confirms the results of many earlier observational studies.

### Secondary outcomes

In general, a trial of CPAP does not reduce overall survival through delay of IMV and may be safely implemented in the absence of criteria indicating an immediate need for IMV such as reduced consciousness or hypotension. NIV may possibly confer a survival benefit in a certain group of patients failing CPAP.

However, there are significant gaps in our understanding, particularly regarding optimal CPAP protocols, definitions of CPAP failure and any possible added benefit from NIV. Economic analyses for CPAP in CARDS are also needed to support public health policy. While Covid 19 ARDS is thankfully in the decline, further research in these areas is needed for a robust response to the next viral ARDS pandemic.

## Data Availability

All data produced in the present work are contained in the manuscript

## Supplementary file 1

### Individual study commentaries based on CASP analysis

#### 1. Brusasco et al (2021)(19)

This single centre cohort study conducted in Northern Italy reports a high CPAP success rate (53/64 = 83%), however patients with a Do Not Intubate (DNI) order are excluded from the study. DNI patients are likely to be frailer and more comorbid at baseline, creating bias in sample selection removing patients who would not be expected to do well on CPAP. The mean pre–CPAP P_a_O_2_/FiO_2_ was 119, in keeping moderate to severe ARDS as described in the study. Significant limitations of this study include single centre study and lack of control arm making it difficult to judge whether CPAP had improved care compared with a standard or comparator of simple oxygen before IMV. However, the survival outcome is still good against a comparator of 30 – 60% mortality which is quoted for ARDS.

Despite limitations, overall, this was an important study early in the pandemic when the evidence base for CPAP in CARDS was very limited.

#### 2. Aliberti et al (2020)(20)

This multicentre cohort study also conducted in Northern Italy has a larger sample size than Brusasco et al (n = 157 cf 64) and has the advantage of being conducted across multiple centres. This study also differs from Brusasco et al in including patients with mild CARDS as candidates for helmet CPAP (mean pre–CPAP P_a_O_2_/FiO_2_ is 146 mmHg, cf 119 mmHg). However, while patients with mild disease who would be expected to do better in terms of survival are included, this study also includes a significant number of patients (65/157 = 41% of total) with a DNI status who would have a poorer functional baseline and therefore be expected to do worse. Notably 36/41 (88%) of patients with a DNI status died despite CPAP.

When both DNI and full escalation (non DNI) groups are considered together there is a lower CPAP success rate compared with Brusasco et al (87/157 = 55%, cf 83%). However, it difficult to assess if these differences are significant as 95% confidence intervals are not quoted.

Excluding patients with DNI status, CPAP success rate is much higher at 72%. Further, of the patients intubated after failing CPAP 25/34 (74%) survived. Therefore, among patients without a DNI status, overall survival (CPAP + IMV) is high at 90% (83/92). While this is higher than Brusasco et al, it includes patients with mild disease who would have been expected to better in any case.

Also, criteria thresholds for CPAP failure differ from Brusasco et al, for example a respiratory rate > 25 is employed rather than > 30 as in Brusasco et al. As later studies have shown tachypnea to be a poor discriminator of CPAP failure(25,28), some the patients deemed as CPAP failure in Aliberti et al may have had the potential to recover with IMV.

Finally and interestingly patients with CPAP success showed a greater proportion with significant (>30%) initial improvement of P_a_O_2_/FiO_2_ with CPAP, however many patients with significant initial improvement did die. Even so, improvement on oxygenation(24) or lung recruitability(11) with CPAP has been used as a predictor of CPAP success in other studies.

#### 3. Franco et al. (2020)(21)

This was a larger cohort study (n= 670) with patients allocated helmet CPAP as the non-invasive respiratory support modality of first choice (n = 330), with NIV (n=177) and High Flow Nasal Oxygen (HFNO, n=163) when CPAP was not available. A 70% (230/330) success rate with CPAP was reported. Authors report no significant difference in survival between CPAP and HFNO or NIV. Even so, HFNO was used in less sick patients than CPAP. Also, there is no clear definition of CPAP failure and practices may have varied between centres. The paper concludes that CPAP/HFNC/NIV outside ITU has favourable survival characteristics.

#### 4. Radovanovic et al (2021)(22)

This is important as a systematic review of CPAP success rates and survival outcomes as well as comparing CPAP with NIV. This review includes 23 studies from Medline and Embase which focus specifically on CPAP (n = 1061) and NIV (n = 1011) for CARDS. However, 2 of the 23 studies (n = 90) did not distinguish between CPAP and NIV outcomes and consequently are not included in the overall CPAP and NIV success rates and survival data. Most of the studies 18/23 were from Europe (Italy 8, UK 7), with 3 from China. All were cohort studies by design.

CPAP had a 51.3% success rate and 77.8% survival rate overall. Here notably the failure (58.8% cf 48.7%) and mortality rate (35.1% cf 22.2%) with NIV was higher than with CPAP. However, no confidence intervals for data are presented.

Additional secondary outcomes are presented including length of hospital stay with a median of 18 days for CPAP. Complications of CPAP are under reported, appearing in only 5/23 studies. 2 studies report 1 case of pneumothorax each which may be associated with CPAP barotrauma.

As with previous studies, there is marked heterogeneity in CPAP initiation thresholds, application method, assessment of failure and weaning.

However, this study is still an important synthesis of early Covid 19 data showing the feasibility of CPAP for CARDS across centres in multiple countries.

#### 5. Hoole et al (2022)(23)

This study was conducted in a respiratory support unit at a secondary hospital in a resource poor setting (Northern Pakistan) with no escalation to IMV due to scarcity of availability. All patients meeting moderate to severe Covid 19 ARDS criteria received CPAP regardless premorbid status.

CPAP success rate and survival rate (which were the same given lack of access to IMV) are higher than some other studies(32), although no confidence interval is given. There are several possible reasons for this phenomenon. First, the patient cohort is likely younger than in other European studies, given South Asian regional age demographics (median age in Pakistan is 24 years cf 48 years in the UK), and younger patients were shown to have better outcomes with Covid 19 disease including severe disease such as ARDS. Secondly, there was routine adoption of prone positioning that is lying patients on their front, to improve oxygenation at the back of the lung, where more lung is located. This has been shown to improve CPAP outcomes in other studies. Finally, there was continual employment of CPAP in circumstances where patients in resource rich settings would have been escalated to IMV.

Although significantly limited in being a single centre observational study with no control group, this preliminary study is still useful in showing what can be achieved with limited resources.

#### 6. Coppadoro et al (2021)(24)

This study is similar to Franco et al and included a cohort of patients with mild to severe CARDS to receive helmet CPAP. However overall CPAP success rate is lower at 52%. But success rate is significantly higher at 69% in the full treatment group versus 28% in the DNI group.

A doubling of PaO2/FiO2 ratio was significantly higher (p < 0.05) in the CPAP success group compared with the CPAP failure group, although even among patients with CPAP failure there was a significant improvement in oxygenation (P_a_O_2_/FiO_2_) and respiratory rate with CPAP compared with simple oxygen.

An interesting aspect of limitations mentioned in the study includes the difficulty of adhering to protocols due to increased clinical burden, which is likely true of many critical care studies during the pandemic.

#### 7. Isaac et al (2022)(25)

This single centre cohort study conducted at a tertiary hospital in South India (Christian Medical Centre, Vellore) consisted of patients with moderate CARDS, with a narrow range of FiO2 requirements (0.4 – 0.6) to maintain oxygenation.

Overall CPAP failure success rate is low (41%), although with added NIV the combined success rate of non-invasive respiratory support (NRS) is higher (77%). The combined success rate is interestingly similar to the Hoole et al study (76.8%) also in South Asia with a similar Covid 19 patient demographic (Bach Christian Hospital, KPK in Northern Pakistan) but which did not employ NIV as a step up. Reasons for this high failure rate of CPAP, but similar NRS success rates overall are unclear. These may include lower threshold criteria for CPAP failure, and a lack of trials of higher CPAP pressures (e.g., 15 cmH_2_O, as opposed to just 10 cmH_2_O).

Additionally, escalation to ITU for intubation and ventilation and higher-level NIV care was available at this south Indian institution unlike the one in Pakistan. The availability of escalation to ITU likely contributed to a higher all survival rate for moderate to severe Covid 19 ARDS (83.3%), and for patients failing NIV the probability of survival was 33.3%.

Finally, significant limitations include a lack of objective data on oxygenation such as P_a_O_2_/FiO_2_ ratio which are routinely reported in other studies and help gage the effectiveness of changes in respiratory support(19,23). Further data on the type of respiratory failure would also have been useful to judge the effectiveness of NIV over CPAP alone. However, this still remains a significant study on CPAP in CARDS in a Lower Middle-Income Country (LMIC) context.

#### 8. Coppola et al. (2021)(26)

This is a similar study to Isaac et al with CPAP first line for CARDS and a NIV trial before IMV for patients failing CPAP. However, patients with severe CARDS are still enrolled in the study, but notably patients with DNI orders (43/199) are excluded. CPAP success rate is 60% (93/156) with an additional 10% success with escalation to NIV (70% total, 109/156). Detailed P_a_O_2_/FiO_2_ metrics are given in this study which is useful.

However notably in this study (and in Isaac et al) – CPAP pressures were limited to 10 cmH_2_O, as opposed to Hoole et al where CPAP pressures up to 15 cmH_2_O were used. Given broadly similar CPAP/NIV success rates in these three studies it is unclear whether the NIV truly has an added advantage to CPAP, or whether similar outcomes could be achieved by utilising higher CPAP pressures (up to 15 cmH_2_O in Hoole et al) in patients failing lower pressures (i.e., 10 cmH_2_O).

#### 9. Ashish et al (2021)(27)

This is a very preliminary study from the UK in which only 18 patients received CPAP (3/103 in the late CPAP group vs 15/103 in the early CPAP group). 1/3 patients on CPAP survived in the late group, compared with 8/15 in the late group. The study concludes that early CPAP was associated with better survival. However, the size of the samples is so small and given the retrospective observational methodology, it is difficult to conclude much from this study with confidence.

#### 10. Wozniak et al (2022)(28)

This study was a retrospective cohort study conducted at a well-resourced tertiary centre. The sample size of CPAP patients is small (23) but all patients had been referred from neighbouring hospitals for consideration of IMV and most had severe CARDS (mean P_a_O_2_/FiO_2_ 84.3 +/- 19 mmHg). The CPAP success rate is 61% (14/23) despite severe CARDS where other centres may have considered IMV. All patients (9/9) who failed CPAP survived with IMV, suggesting that a trial of CPAP in these patients did not worsen outcomes through delaying IMV. However, patients were monitored well and most patients failing CPAP were intubated early within 6 hours CPAP initiation. Further this cohort likely had a good physiological baseline, given eligibility for transfer to a specialist ICU at a time of resource scarcity, and the results of this trial are less likely to the broader population containing more elderly, frail, and comorbid patients.

All patients in the study showed improvement in P_a_O_2_/FiO_2_ with CPAP, with mean P_a_O_2_/FiO_2_ almost doubling. However, there was not a significant difference between CPAP responders and non-responders, unlike Coppadoro et al. Interestingly CPAP also did not significantly decrease respiratory rate either in CPAP responders or non-responders, suggesting that change in respiratory rate may be a poor discriminator of CPAP success. However, d dimer, CRP and IL 6 were significantly more elevated in patients who failed CPAP, than those who succeeded. Additionally, 5/6 of CPAP non responders who received a CT Pulmonary Angiogram (CTPA) to detect pulmonary embolism (PE) were found to have PE, suggesting that additional factors such as PE may influence response or non-response to CPAP in CARDS.

While the findings may have limited generalisability to other settings due to the highly selective small sample size and high resource setting in which this study was conducted, the study remains significant due to a high CPAP success rate despite severe Covid 19 ARDS.

#### 11. Perkins et al (2022) – Recovery RS Trial (29)

This is one of the largest Randomised Control Trials (RCTs) on CPAP and HFNO vs conventional oxygen (N = 1273). Intubation free survival at 30 days was significantly greater in the CPAP group (240/377 = 64%) compared with simple oxygenation (198/356 = 56%), but not in HFNO (231/415 = 56%) compared with simple oxygen. The trial recruited less than half of the number of patients initially intended due to declining cases of severe Covid 19 towards the end of the pandemic. Nonetheless this was a landmark study that shifted National Institute of Clinical Excellence (NICE) guidance in favour of CPAP in Covid 19 ARDS in patients with an FiO2 requirement of 0.4 or greater, without need of immediate (within 1h) intubation(13).

Serious adverse events related to CPAP include 3 patients with barotrauma (pneumothorax or pneumomediastinum).

Limitations include significant crossover between groups, inability to blind participants and clinicians to the intervention and possibly insufficient power to detect difference between HFNO and simple oxygen.

#### 12. Vaschetto et al (2020)(30)

A large retrospective cohort study with CPAP survival rates broadly similar to other larger studies with a 55% CPAP success rate, with an overall 66% survival (29% in the full escalation group and 27% in the DNI group). Delayed intubation noted to be a risk factor for mortality, but without controls it is difficult to interpret as intubation may have been delayed in patients with a lesser chance of overall survival given limited resources.

#### 13. Lawton et al (2021)(31)

This is an early single centre retrospective cohort study with a 62% intubation free survival in the full escalation group in a comorbid population. Mortality in the CPAP group with DNI status was 70%. Early CPAP and routine prone positioning were adopted.

